# A Bayesian approach to identifying the role of hospital structure and staff interactions in nosocomial transmission of SARS-CoV-2

**DOI:** 10.1101/2023.09.11.23295353

**Authors:** Jessica R.E. Bridgen, Joseph M. Lewis, Stacy Todd, Miriam Taegtmeyer, Jonathan M. Read, Chris P. Jewell

## Abstract

Nosocomial infections threaten patient safety, and were widely reported during the COVID-19 pandemic. Effective hospital infection control requires a detailed understanding of the role of different transmission pathways, yet these are poorly quantified. Using patient and staff data from a large UK hospital we demonstrate a method to infer unobserved epidemiological event times efficiently and disentangle the infectious pressure dynamics by ward. A stochastic individual-level, continuous-time state-transition model was constructed to model transmission of SARS-CoV-2, incorporating a dynamic staff-patient contact network as time-varying parameters. A Metropolis-Hastings MCMC algorithm was used to estimate transmission rate parameters associated with each possible source of infection, and the unobserved infection and recovery times. We found that the total infectious pressure exerted on an individual in a ward varied over time, as did the primary source of transmission. There was marked heterogeneity between wards; each ward experienced unique infectious pressure over time. Hospital infection control should consider the role of between-ward movement of staff as a key infectious source of nosocomial infection for SARS-CoV-2. With further development, this method could be implemented routinely for real-time monitoring of nosocomial transmission and to evaluate interventions.

## Introduction

Healthcare-associated infections are a significant burden to health systems worldwide, and are associated with increased morbidity and mortality [1]. Transmission of SARS-CoV-2 in healthcare settings was widely reported during the first wave of the COVID-19 pandemic [2–5]. Hospital patients are considered to be especially vulnerable to severe COVID-19 and healthcare workers have been shown to be at an increased risk of infection [6–8]. Identifying nosocomial transmission routes of SARS-CoV-2 is therefore critical to patient and staff safety. Universal point of care testing of patients and staff was not available during the initial stages of the pandemic in the UK to rapidly identify and isolate individuals. A triage system was recommended in hospitals to isolate and test patients with suspected COVID-19 [9]. Although hospitals employed a range of infection prevention control (IPC) protocols to minimise transmission, high levels of hospital infections were reported [2, 10].

To determine nosocomial transmission routes of a respiratory pathogen, the network of transmission opportunities within the hospital, hereafter called the contact network, must be considered. The routine running of a hospital in the UK, comprising of wards, bays and side rooms, can give rise to additional structural and network transmission opportunities. Partitioning of patients into wards may be by diagnosis, severity of illness, or by infection status during an outbreak. Patients may have direct contact with staff, visitors and patients in the same ward, but also indirect contact with other patients and staff through shared equipment and objects, relevant to fomite transmission, airflow, relevant to airborne transmission, and staff acting as vectors for infectious diseases. Modelling studies have been conducted to investigate nosocomial transmission routes of numerous pathogens, most commonly *Methicillin-resistant Staphylococcus aureus* (MRSA) and *Vancomycin-resistant Enterococcus* (VRE) [11]. While transmission from patients to patients and healthcare workers to patients has been identified, there remains a lack of consensus as to the primary routes of nosocomial infection [7, 12, 13].

One of the challenges of identifying transmission routes in a hospital is distinguishing between hospital-acquired and community-acquired infections. Knight et al. report large uncertainty in the classification of nosocomial and community-acquired infections of SARS-CoV-2 during the first part of 2020 [14]. Routinely collected hospital data tends to record a timestamp of when a patient tested positive for an infection. For COVID-19, this provides a marker as to when a patient was infectious. However, the time that a patient became infected is unobserved, and the time a patient recovered from infection and ceased to be infectious is also often unobserved. This makes the inference for nosocomial models much more difficult and computationally intensive, as these unobserved epidemiological events need to be mapped on to the structured landscape given by the composition of the hospital.

Generally, surveillance data only captures a partially observed epidemic process, and therefore requires a form of data augmentation to estimate unobserved epidemiological event times. O’Neill et al. initially introduced a Bayesian data augmentation approach to inference of general stochastic epidemic models, where unobserved event times are treated as parameters to be estimated [15]. This has proved to be a popular method for conducting inference on partially observed epidemic models [16, 17]. A drawback of this method is that repeatedly calculating the likelihood can become extremely costly computationally and its use is therefore limited by population size. As an alternative method, McKinley et al. demonstrate using approximate likelihood ratios to conduct inference on partially observed epidemics, though this relies on repeatedly simulating the epidemic [18]. Previous studies which model nosocomial transmission have used data augmentation to handle unobserved infection times [19, 20]. However, these studies do not include a time-varying contact network parameter to model interactions between staff and patients.

Here we use routinely collected data from a large acute-care hospital in the UK to quantify the temporal and network dynamics of nosocomial transmission of SARS-CoV-2. We demonstrate a Bayesian approach to conducting inference with a time-varying covariate, a fine-scale patient-staff contact network, to estimate unobserved epidemiological event times and provide insight in to within-hospital transmission dynamics.

## Covariate Data

Patient testing data from a large acute-care hospital in the UK was used in conjunction with staff rota data to identify routes of nosocomial transmission of SARS-CoV-2 during the first wave of the COVID-19 pandemic. A patient pathway was developed with a colour coded ward system to separate patients based on their SARS-CoV-2 infection status; Figure 1. Universal testing for SARS-CoV-2 was not available for patient admissions or staff during our study period. Patient diagnosis was therefore based on clinical suspicion of COVID-19 and a confirmatory test. Some wards (e.g. the isolation ward) could be assigned different colour areas within a single ward based on the availability of side rooms (enclosed patient rooms), which might be treated differently than communal areas. Ward colours could also change over time in view of capacity demands. PPE guidance for staff differed by ward colour. Our study period covered a four week time span from 12 April 2020 to 10 May 2020. During this time, 3,816 staff worked at least one shift at the hospital, consisting of 2,948 healthcare staff, 232 medical doctors and 636 ancillary staff. There were N = 2,981 patients admitted (including day attenders) in our study period to p = 55 wards.

**Figure 1:**
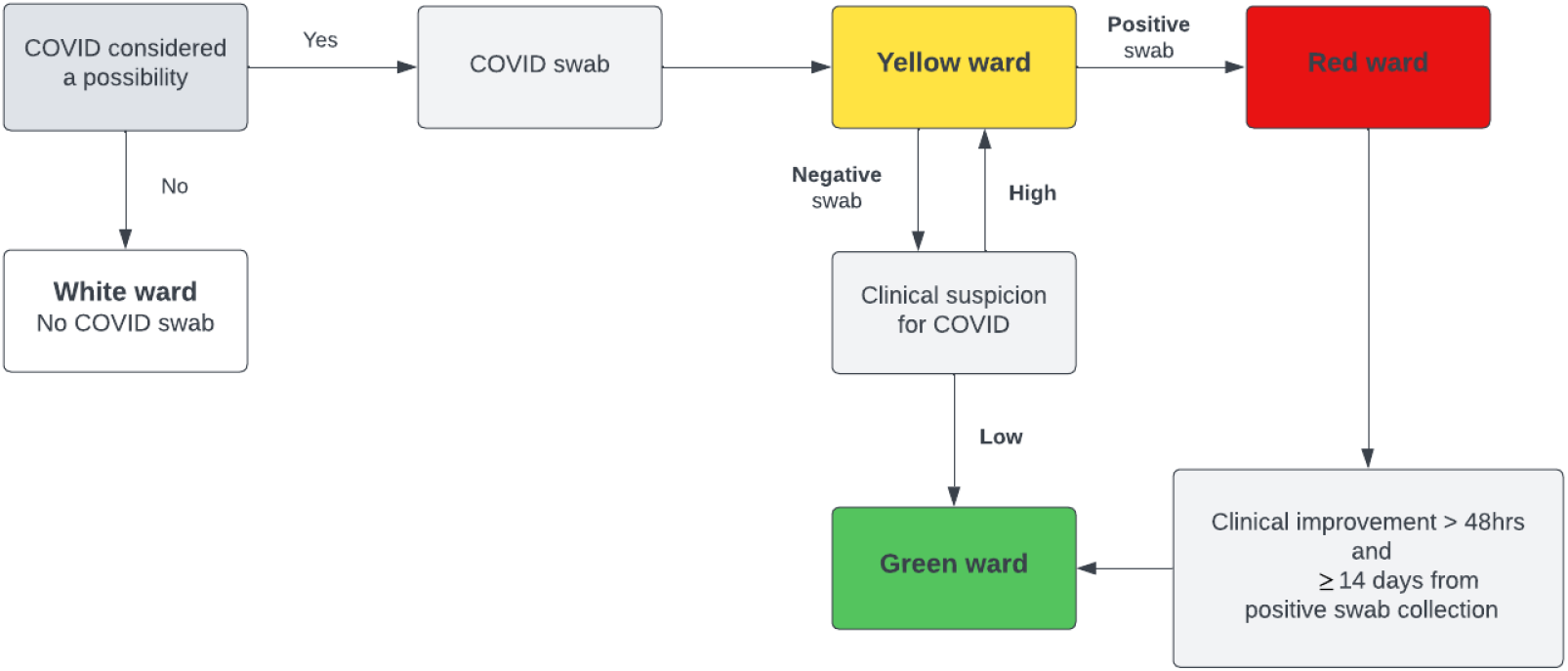
Patient pathway from initial admission to ward allocation, by suspected and confirmed SARS-CoV-2 infection status, during the study period.

Anonymised patient location, staff work patterns and SARS-CoV-2 testing results were extracted from routine trust electronic records. The study received HRA approval via IRAS and trust approval as a research project, accessing only routinely collected anonymised data (IRAS ID 288257).

### Hospital Network

Patient movements (ward transfers, admission and discharges) and staff shift times were recorded continuously in time. However, the recorded time of these movements were not likely to be exact in practice, and the volume of changes to the patient and staff structure leads to prohibitively large data structures in RAM. Hence, patient and staff movements were aggregated to one hour time intervals to form the basis of our discrete-time contact network.

The hospital contact network is represented in three distinct ways. Firstly, a weighted ward connectivity tensor, *C*, of shape [*p* × *p* × *T*], where element *c*_*qrt*_ is the connectivity between ward *q* and *r* at time *t*, in units of number of staff working across multiple wards. This primarily consists of doctors who are assigned to a group of wards per shift. Similarly, we define a spatial adjacency tensor of shape [*p × p × T*] denoted *W*, where *w*_*qrt*_ *>* 0 if wards *q* and *r* share a kitchen, and zero otherwise. Given the wards do share a kitchen, *w*_*qrt*_ represents the number of staff allocated the use of that kitchen at time *t*. Figure 2 illustrates how wards may be connected by kitchens, albeit separated by storey. Lastly, a membership tensor of shape [*N × p × T*] denoted *M*. Where *m*_*iqt*_ = 1 if individual *i* is a member of ward *q* at time *t* and zero otherwise. Visualisations of the contact network can be found in the supplementary material.

**Figure 2:**
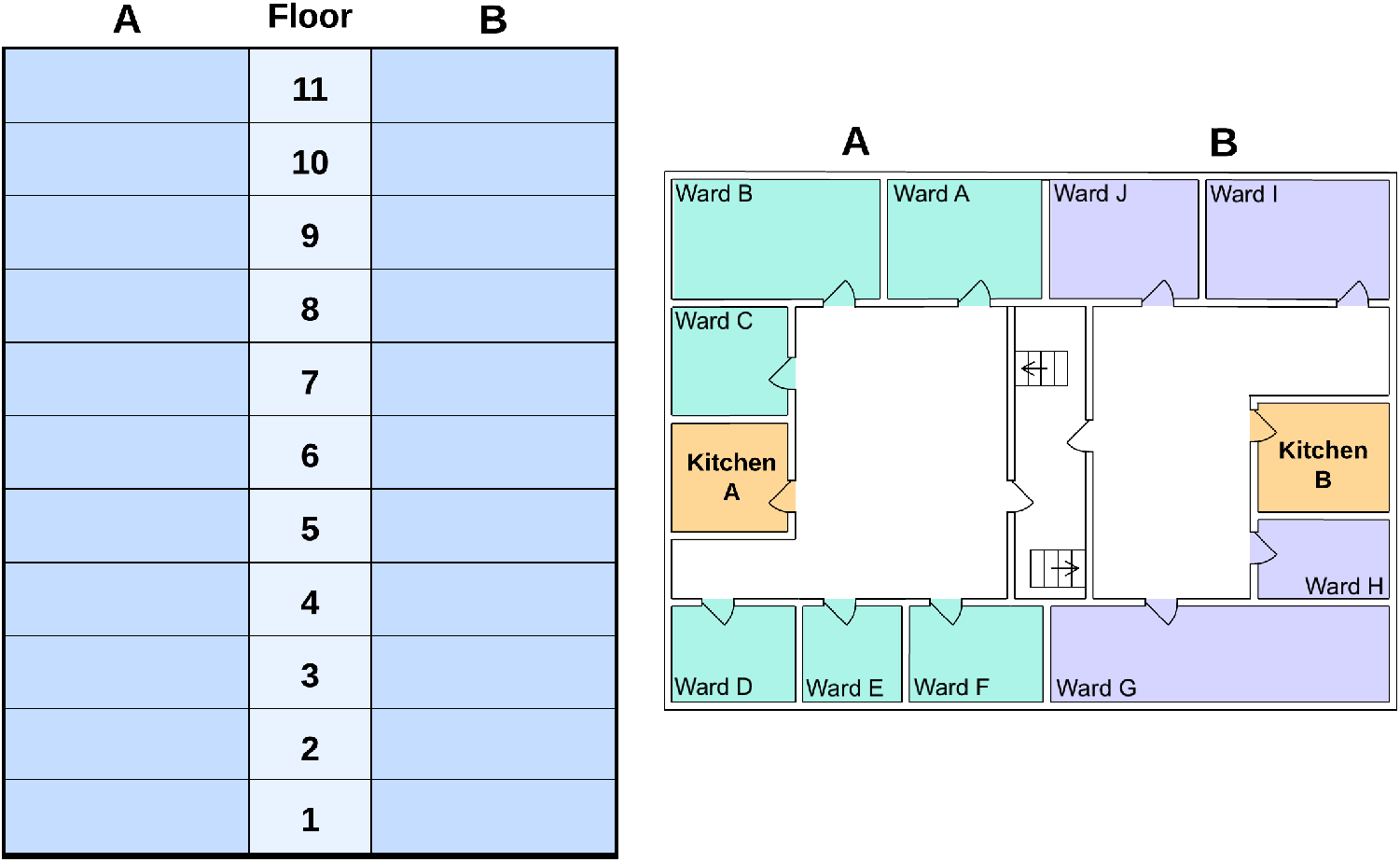
A schematic illustration of the hospital layout. Each floor of the hospital has two kitchens, one on side A and one on side B. The spatial adjacency tensor W, as defined previously, would consider all wards on the the same floor and side of the hospital as connected.

We introduce the dot subscript notation to indicate a slice of a tensor. For example, *C*_*q·t*_ refers to a slice of tensor C along the second axis, which can be thought of as a vector representing the connectivity between ward *q* and all hospital wards (*∀r*) at time *t*.

## Modelling

### Model Structure

Transmission of SARS-CoV-2 is represented here by a stochastic individual-level, continuous-time state-transition model. At any timepoint, patients are considered to belong to one of four mutually exclusive epidemiological states: *susceptible to infection (*S*), infected but not infectious (*E*), infected and infectious (*I), or *recovered/removed from the population (R)*. Any particular patient, *i* is assumed to progress between the states according to the transitions [SE], [EI], and [IR] with transition rates 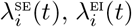, and 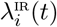 as defined below.

At time *t*, the rate at which patient *i* = 1, …, *N* becomes infected, i.e. 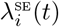, is assumed to be a function of the time-evolving infectious landscape around them. Within the hospital, we assume that *i* experiences infection pressure from four separate sources: other patients in the same ward; patients in other wards connected by staff being assigned to more than one ward; the spatial structure of wards connected by adjoining kitchens; constant ‘background’ infection, representing sources of infection not explicitly modelled by the hospital structure. Within-hospital routes of infection are represented by our patient-staff contact networks (*C, W, M*). Within the continuous-time model, changes to the patient-ward and staff-ward structures (either via patient movements or staff-ward allocation) are assumed to occur at discrete intervals of one hour, aggregating the precisely time-stamped patient movement and staff shift data to the hour. Continuous-time was used to model the epidemic process to account for the the fluidity of events in the hospital through time, thus avoiding the necessary act of choosing a time-step. In addition, this leads to more efficient sampling from the posterior distribution of censored event times, avoiding large swings in posterior density that otherwise occur in discrete-time systems.

The rate at which an individual transitions from state S to E can be described as a time-dependent infectious pressure. The infectious pressure is density dependent and is defined at the individual-level. We account for the number of infected on each ward at time *t*, the patient-staff contact network at time *t* and a background infection rate. An individual can either be present in the hospital *H* or in the community. The infectious pressure on an individual *i* in ward *q* at time *t* can be defined as:

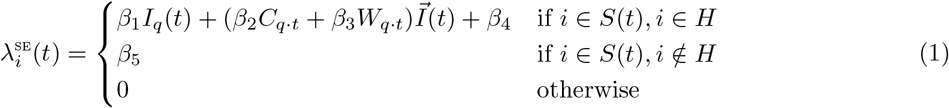

where *β*_1_ denotes the transmission rate for within-ward mixing and *I*_*q*_(*t*) represents the number of infected individuals on ward *q* at time *t. β*_2_ and *β*_3_ are transmission rates for between-ward mixing, with *C*_*q·t*_ denoting the connectivity between ward *q* and all other wards at time *t*, and *W*_*q·t*_ describing the connectivity between ward *q* and all other wards by ward proximity at time *t*. 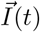 represents the number of infected individuals on each ward and *β*_4_ is a background hospital transmission rate. To allow for individuals to be infected before and between hospital admissions, we define a constant infectious pressure *β*_5_ that is exerted on to susceptibles in the community.

Similarly, we can define an individual’s [EI] transition rate as follows:

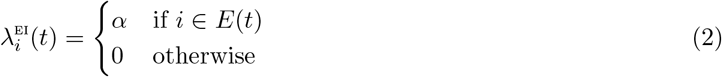

Likewise, an individual’s [IR] transition rate is defined as:

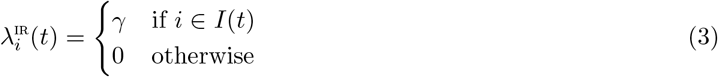

Thus the [EI] and [IR] transition rates are assumed to be constant across individuals and time, and for identifiability reasons we fix *α* = 1*/*4 day^-1^ and *γ* = 1*/*5 day^-1^ respectively [21].

The epidemic process is assumed to be Markovian. The data generating process is outlined in Algorithm 1.

#### Algorithm 1: Gillespie’s Direct algorithm

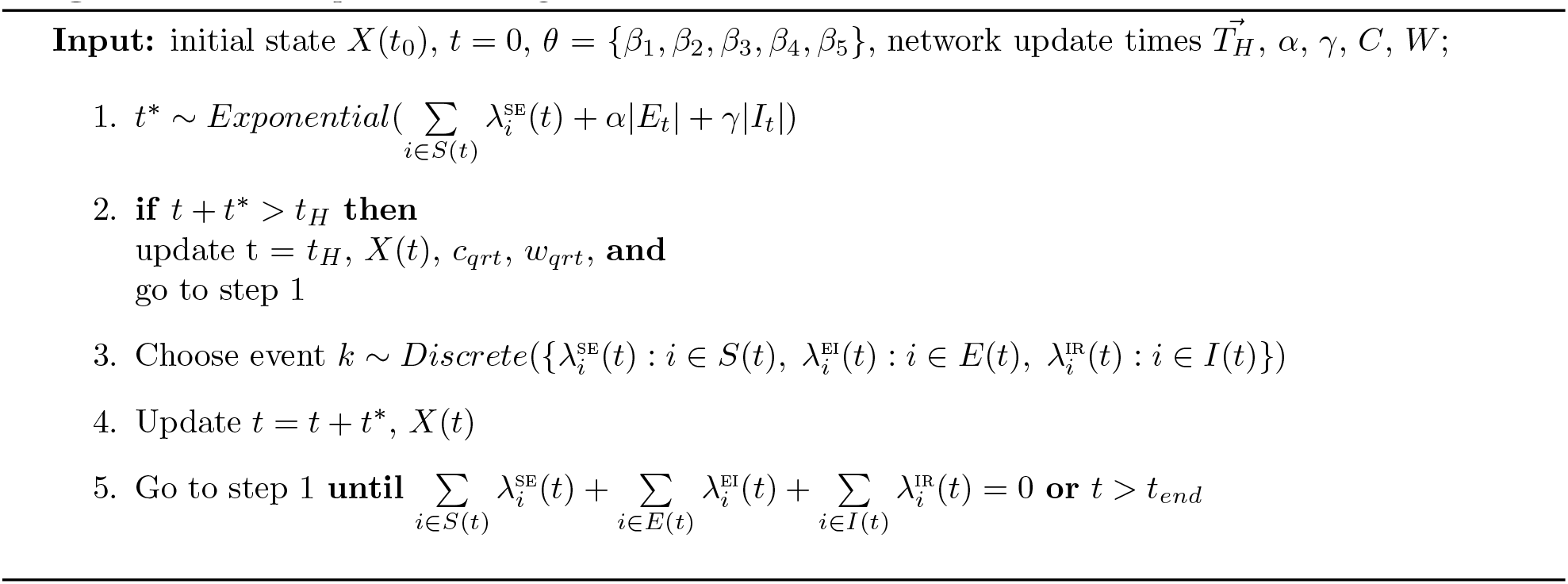

### Bayesian Inference

A Metropolis-Hastings Markov chain Monte Carlo (MCMC) algorithm is used to estimate the transmission rate parameters *θ* = {*β*_1_, *β*_2_, *β*_3_, *β*_4_, *β*_5_} and the unobserved [SE] and [IR] state transition times for each infection event, denoted *t*_*SE*_ and *t*_*IR*_ respectively, according to the method of Jewell et al. [17]. Each infection transition event is associated with an observed [EI] transition time, where an individual’s [EI] transition time is assumed to be two days before the collection of their first positive swab. We assume that infections occur at points of a right-continuous time inhomogeneous Poisson process at a rate equal to the sum of infectious pressure on susceptibles immediately before that time point. The inclusion of a time-varying covariate, the contact networks, in the continuous-time model adds to the complexity of computing the likelihood. We define the likelihood of observing *z* transitions, in terms of an ordered event list, where an event is considered to be a transition event (transitioning between states: S*→* E, E*→* I or I*→* R) or a hospital network update, as follows:

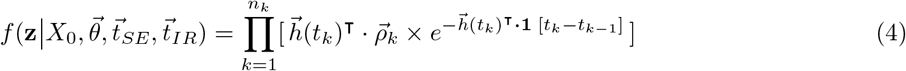

where *X*_0_ denotes the initial conditions, *k* refers to the event index and 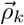 denotes one-hot encoding of the event. 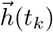 is a vector of length LN+1 of hazard rates for L = 3 transitions, N = 2,981 individuals and a covariate marker to indicate a hospital network update. For example:

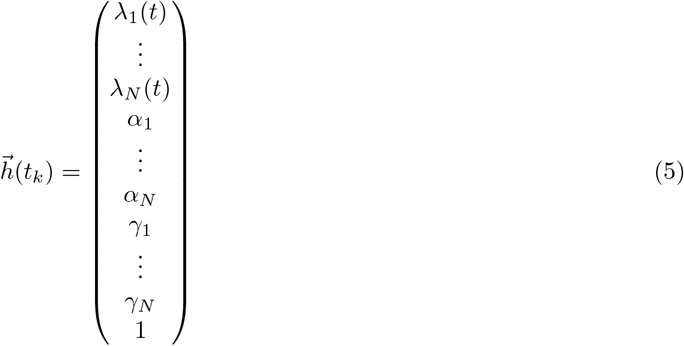

Gamma priors were chosen for all transmission parameters (*β*_1_, *β*_2_, *β*_3_, *β*_4_, *β*_5_) denoted by *f*(*θ*). The joint conditional posterior can therefore be defined as:

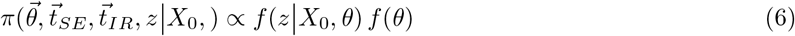

As the likelihood is intractable to integration, a Metropolis-within-Gibbs MCMC algorithm is used to sample from the joint posterior. Full details of the MCMC algorithm used can be found in the supplementary materials. To evaluate chain convergence, the Gelman and Rubin potential scale reduction statistic is calculated for three chains with differing starting values drawn from the prior distribution [22]. The posterior predictive distribution is analysed visually to assess how well our modelled estimates fit the observed data.

### Code Implementation

The model and MCMC were implemented in Python V.3.9 using TensorFlow and TensorFlow Probability for GPU acceleration [23, 24]. The model code is available at: https://github.com/jbridgen/nosocomial_covid_model.

### Infection Hazard Attributable Fraction

Having computed the joint posterior distribution, we are able to investigate how within-ward and between-ward dynamics contribute to nosocomial transmission. We calculate the attributable fraction, denoted *AF*, per infection for each transmission type; within-ward transmission *β*_1_; between-ward transmission rates *β*_2_ and *β*_3_; background hospital transmission rate *β*_4_; community transmission rate *β*_5_. For example, the attributable fraction for individual *i* infected from between-ward transmission on ward *q* can be defined as follows:

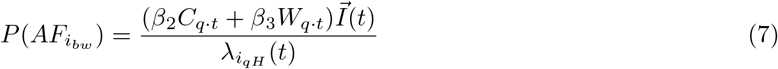

The attributable fractions are aggregated for each set of sampled parameter estimates to compute the mean attributable fraction and 95% Credible Intervals (95%CrI) for each transmission type per infection. Similarly, using the sampled parameter estimates we can identify which wards and associated ward colours nosocomial infections take place in.

## Results

The results presented are of SARS-CoV-2 infections recorded in a UK hospital one month in to the first wave of the pandemic. In a population size of 2,981 patients from 12 April 2020 to 10 May 2020, we have identified 131 infection events, [EI] transitions, which are a combination of community-acquired and nosocomially-acquired infections. Historic testing data, 3 days prior to our study start date, was used to identify the 58 patients which are considered to be initially infectious, residing in the I state, in our model.

### Parameter Estimation

To estimate our parameters of interest (*β*_1_, *β*_2_, *β*_3_, *β*_4_, *β*_5_, *t*_*SE*_, *t*_*IR*_), we ran a Metropolis-Hastings MCMC algorithm for 11,000 iterations removing the first 1000 samples as burn-in. Convergence across all parameters is seen which is confirmed by the potential scale reduction statistic computed for three independent chains; see supplementary materials. In Figure 3, we present the kernel density estimates for each of the transmission rate parameters and for a random sample of transition event times. A constraint when drawing event times, is that an individual must have been admitted to hospital before their [IR] transition time. As the observed [EI] transition times are set to two days prior to a patient’s first positive hospital test, there may be a period of time pre-admission which is unexplored by the MCMC algorithm, as seen with sample 1 and 6 of Figure 3B.

**Figure 3:**
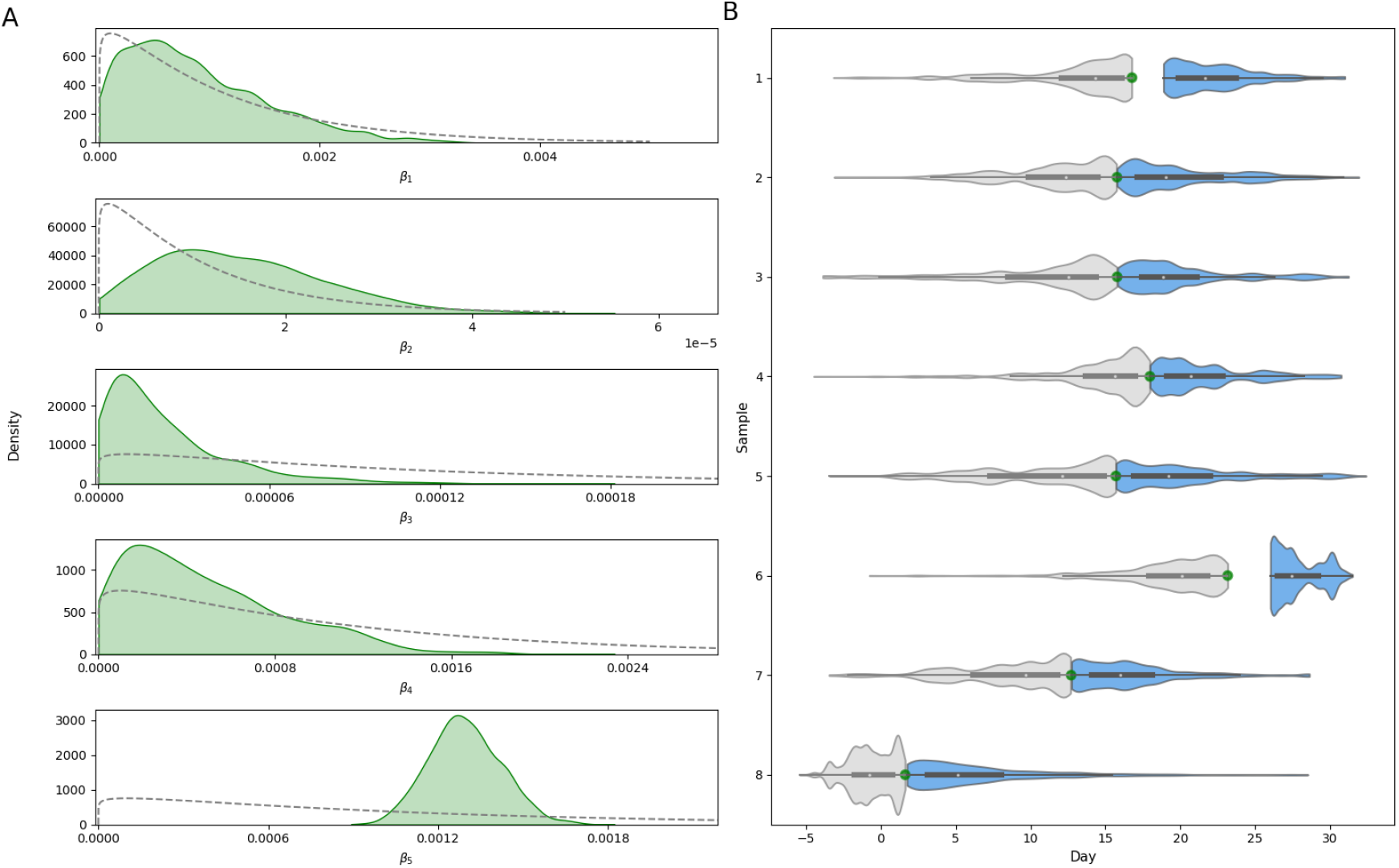
(A) Kernel density estimates for each transmission rate parameter. Dashed lines represent the associated prior distribution.(B) Density estimates of [SE] and [IR] transition times for a random sample of eight observed [EI] events. Green points represent the observed [EI] transition time. Distributions for the associated [SE] and [IR] transition times are shown in grey and blue respectively.

The posterior predictive distribution is used to evaluate how well our model fits the observed data. An epidemic is simulated forward using the data generating model (Algorithm 1) for each of the 10,000 sets of parameter estimates sampled by the MCMC algorithm. We compare the number of [EI] transitions occurring each day from the simulations with the observed data; Figure 4. With the exception of one peak of [EI] transitions on day 12, the observed data sits comfortably within the 95%CrI of the aggregated simulation data. Figure 4 shows that in a given simulation, the peaks and troughs of the number of [EI] transitions per day may similarly sit outside of the credible interval.

**Figure 4:**
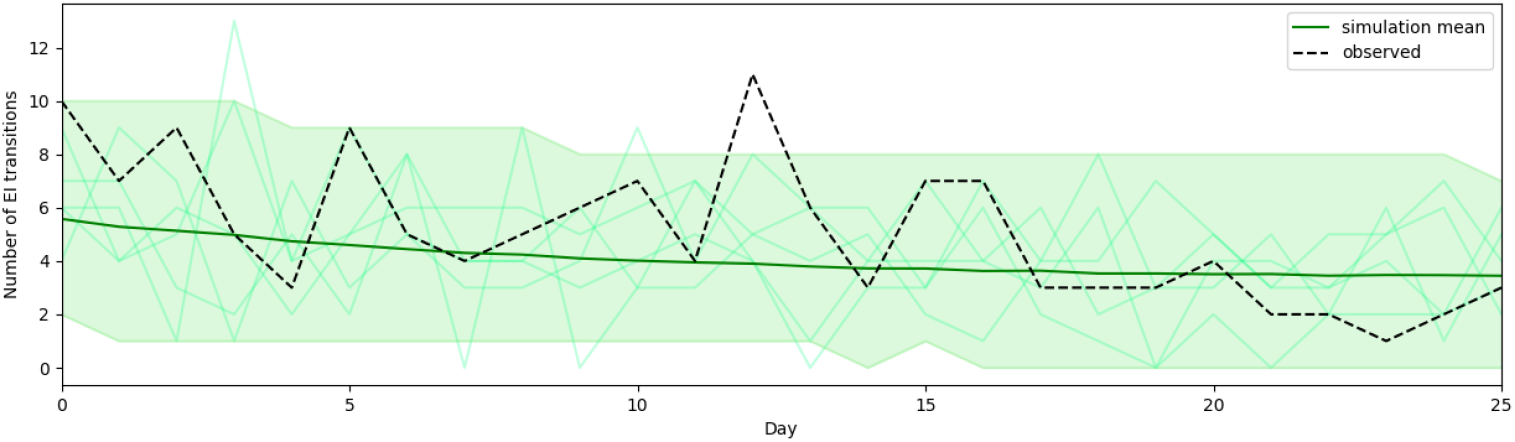
Posterior predictive formed from 10,000 stochastic simulations over the joint posterior. Mean simulated number of [EI] transitions in dark green with 95% Credible Interval as the shaded area. Five individual simulations are displayed in the faint green lines. The observed number of [EI] transitions per day is shown by the dashed line.

### Nosocomial Transmission Routes

One of the parameters of interest is the associated [SE] transition time for each infection. Estimating this enables us to identify which infections were most likely to be contracted nosocomially rather than in the community. Moreover, for each infection we analyse how the different types of transmission contribute to the infectious pressure exerted on an individual directly before their infection event. Hospital-acquired infections are likely for 15.3% (20/131) of detected infections. These infections have a mean community attributable fraction of less than 0.5, with the majority (15/20) having a mean community attributable fraction of less than 0.1; Figure 5. Between-ward transmission is the highest mean attributable fraction for 13 of the 20 infections identified as nosocomial. Infection events 4 and 15 have the highest mean attributable fractions for between-ward transmission of 0.72 (95%CrI 0.12 to 0.99) and 0.83 (95%CrI 0.29 to 1.00); Figure 5. Four nosocomial infections have a mean within-ward attributable fraction of zero, indicating that these infections occurred when there were no infectious individuals admitted to the individual’s ward.

**Figure 5:**
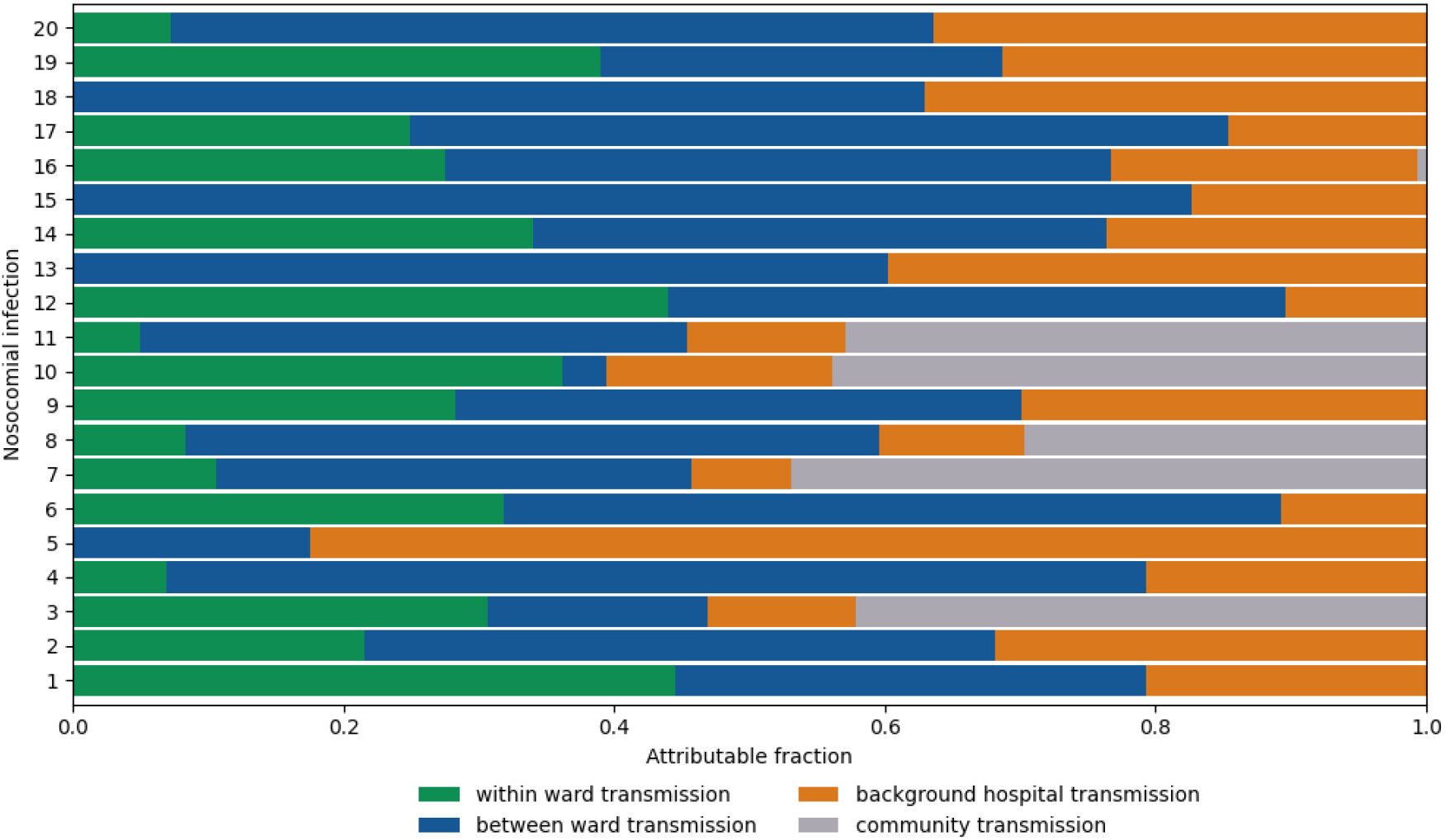
Mean attributable fraction for each transmission type per infection event for 10,000 posterior samples. Infection events are displayed if they have a mean community transmission attributable fraction less than 0.5

To assess the plausibility of our model output, we further explored the hospital data associated with the 15 individuals that were identified as most likely to have hospital-acquired infections. One individual appeared to have been tested on the day they were admitted to hospital; on further inspection, this individual had been discharged from a 15 day hospital spell two days prior to readmission which was captured within our study period. The other 14 individuals identified had been admitted to hospital for an average of 13.3 days (95%CrI 6.62 to 22.76) before their positive test sample was collected.

Using our modelling framework we are able to identify the wards in which nosocomial infections likely occur. Nosocomial infections are identified based on whether an individual’s [SE] transition time is during a hospital admission spell. For each set of sampled parameters, the percentage distribution of nosocomial infections by ward is calculated and aggregated for the 10,000 samples. We find that four wards account for the locations of 63.1% of nosocomially-acquired infections, with wards 13 and 23 accounting for 21.1% (95%CrI 11.76 to 30.00) and 17.5% (95%CrI 10.00 to 25.00) respectively. Similarly, we compute the percentage distribution of nosocomial infections by ward type; Table 1. Nosocomial infections are recorded most frequently in green wards (40.0%, 95%CrI 33.33 to 60.00) and wards which are assigned three colours of green, red and yellow (13.6%, 95%CrI 6.67 to 25.00).

**Table 1:** Percentage of total nosocomial infections by designated ward colour.

| Ward colour | Percentage of nosocomial infections (95%CrI) |
| --- | --- |
| green | 40.04 (33.33-60.00) |
| green-red-yellow | 13.59 (6.67-25.00) |
| white | 10.29 (5.56-23.53) |
| yellow | 10.00 (5.56-20.00) |
| white-yellow | 9.20 (5.56-20.00) |
| red | 5.68 (5.00-9.09) |
| green-red | 5.61 (5.00-9.09) |
| red-yellow | 5.61 (5.26-9.09) |

The modelling suggests that the infectious pressure exerted on an individual in a ward changes considerably over time, this may be explained by the dynamic contact network. The within-ward infectious pressure will increase with the number of infectious individuals on the ward. Similarly, if there is an increase of infectious individuals on wards considered to be connected, the between-ward infectious pressure will increase. Interestingly, the source of infectious pressure exerted on individuals in the four wards which recorded the highest number of nosocomial infections was also dynamic; Figure 6. For the first 23 days of our study period, each of these four wards were classified as green wards with the exception of ward 23 which was a mixed ward, either classified as green-red-yellow or green-red ward. We find infectious pressure dynamics varied by ward and ward colour; Figure 6. An individual on ward 33 would have experienced an infectious pressure driven by between-ward dynamics whereas individuals on wards 13 and 22 would have experienced clear peaks in within-ward infectious pressure.

**Figure 6:**
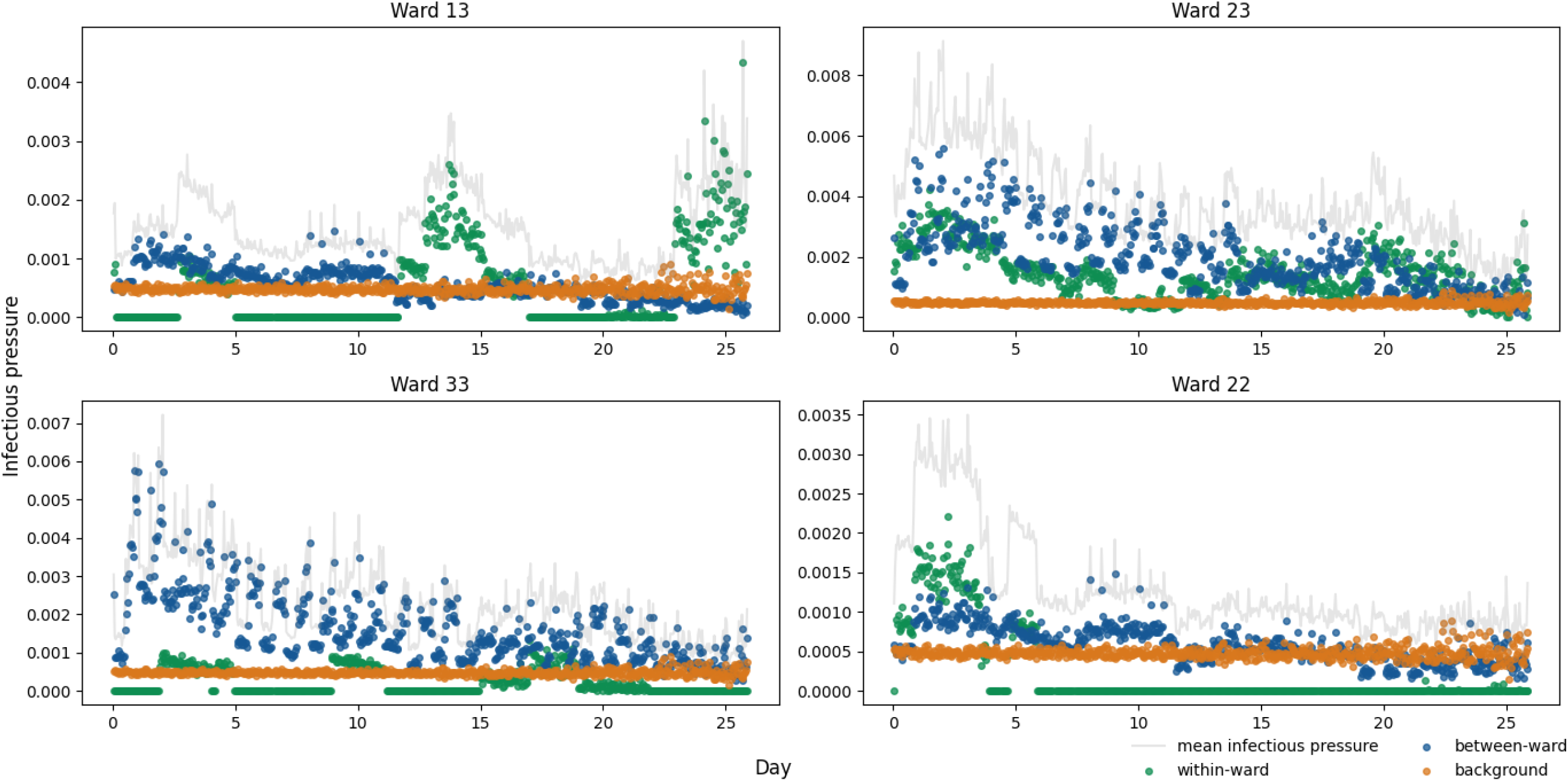
Mean infectious pressure for an individual on the stated ward at each [SE] transition, calculated for 10,000 posterior samples. The wards displayed are those with the highest mean number of nosocomial infections.

## Discussion

This study provides a statistical framework for conducting inference on hospital outbreak dynamics to quantify the relative contribution of transmission routes for nosocomial infections. We have demonstrated that transmission parameters and unobserved event times can be inferred by incorporating a discrete time-varying patient-staff contact network and testing data into a continuous-time stochastic epidemic model. By estimating these parameters we are able to disentangle the different components of infectious pressure and identify routes of nosocomial transmission of SARS-CoV-2 during the first wave of the pandemic. While computing the likelihood for these models can be computationally intensive, we found that with GPU acceleration (1 x NVIDIA V100 32GB), 11,000 iterations of our MCMC took approximately 61 minutes. A Bayesian approach enables us to assess uncertainty in our parameter estimates by simulating forwards stochastically over the joint posterior.

We estimate that 15.3% of identified SARS-CoV-2 infections in patients identified in hospital were nosocomially-acquired, other studies which examined hospital infections during the first wave of the pandemic have reported similar findings [2, 6, 12, 25]. Nonetheless, we expect that this is an underrepresentation of the full extent of within-hospital transmission. Asymptomatic infections are not captured in this study as universal testing of admissions had not yet been introduced. Similarly, if a patient had been discharged while unknowingly infected, their infection would be unrecorded. A simple method which is used to identify nosocomial infections is to consider the time interval between admission and symptom onset. However, this can lead to an infected patient with multiple hospital spells in quick succession being misclassified as a community-acquired infection [14]. The modelling approach presented here enabled us to identify an individual who was discharged from a long hospital stay and re-admitted two days later, as having a likely nosocomially-acquired infection.

We found that the total infectious pressure exerted on an individual in a ward changes over time, as does the primary source of transmission. When comparing wards which housed nosocomial infections, it was clear that infectious pressure dynamics varied greatly by ward. Moreover, these dynamics varied across wards which were designated with the same ward colour. For most nosocomial infections, the most likely source of infection was captured by between-ward dynamics, suggesting that the patient pathway implemented was successful in separating susceptible patients from infectious patients. This finding is supported by several other studies which found that healthcare workers were a likely source of nosocomial infection [13, 26, 27]. Evans et al. found that indirect transmission from infected patients was the most likely route for nosocomial transmission, where indirect transmission may happen through healthcare workers acting as vectors for transmission or through fomite transmission [7]. Without staff infection data it is difficult to pinpoint the exact cause of between-ward transmission. Between-ward transmission in our model is driven by the staff-patient contact network, which indicates that the hospital contact network is a key route for nosocomial transmission.

Nosocomial infections were most likely to be contracted within four wards. These wards tended to be wards designated as green on the patient pathway. Patients would be placed in green wards after receiving a negative test result for SARS-CoV-2 and if they were also of low clinical suspicion of having COVID-19, or if they were being stepped down from a red ward. Separating patients who were not suspected of having COVID-19 on admission appears to have been effective, with fewer nosocomial infections occurring on white wards than green wards. However, undetected asymptomatic transmission may be more likely to occur on white wards as patients were not tested on admission.

There are several limitations to our approach. Firstly, in order to conduct the inference we fix an individual’s [EI] transition time to be two days prior to their first positive test. This allows for a delay between patients becoming infectious and being tested [28]. We do not consider reinfections due to the short length of our study period. Additionally, we assume that individuals progress from exposed to infectious and infectious to recovered at constant rates. As our study period is during the initial phase of the pandemic, vaccination status and virus variants were not considered. The data augmentation methodology used could be developed further to better cope with unknown disease status on admission and occult infections at the end of the time window. Furthermore, the force of infection exerted on the community could be based on prevalence estimates. Community prevalence was not well known for our study period, with estimates often based on hospital admission data, as SARS-CoV-2 tests were not readily available to the public. Nevertheless, this should be considered for future studies. We also did not explore different model structures, using the standard SEIR structure often used for COVID-19 models. Computationally, calculating the likelihood in continuous-time is expensive, it would be worthwhile to investigate the potential gains and losses of a fully discrete model.

Our findings allow us to draw some important conclusions regarding effectiveness of infection prevention and control measures in this hospital early in the pandemic. At this time, universal SARS-CoV-2 testing was not logistically possible, and a lack of isolation facilities meant that patients had to be cohorted based on COVID-19 risk. Firstly, we conclude that stratifying patient risk of SARS-CoV-2 infection based on clinical assessment (primarily presence of respiratory symptoms) was successful, in that few nosocomial infections likely occurred in white (low risk) and yellow (possible COVID-19) wards. Secondly, most nosocomial infections likely occurred in green wards, and was primarily driven by between-ward transmission and hence the staff-patient contact network. These findings can inform future planning for outbreaks of respiratory pathogen and provide support for strategies to reduce staff-patient transmission, such as asymptomatic staff testing and lateral flow testing of patients on admission which were implemented in the NHS later in the pandemic [29].

We have presented a Bayesian approach to quantifying routes of nosocomial transmission of SARS-CoV-2 which could be applied to other respiratory infections and extended to outbreaks of bacterial infections. To our knowledge, inference methods such as the ones presented here have not been used for real-time IPC monitoring due to the computational complexity and impractical runtimes. We have demonstrated an efficient method to infer epidemiological event times and nosocomial transmission routes while accounting for the intricacies of the hospital network. This could be used to retrospectively evaluate and simulate interventions. Additionally, with further development and the appropriate infrastructure in place, we believe that methods such as these could be implemented at a similar scale during a prolonged hospital outbreak to alert IPC teams to potential hotspots of transmission and assess effectiveness of interventions.

## Data Availability

Full model code is available at https://github.com/jbridgen/nosocomial_covid_model. Anonymised patient and staff data are not publicly available due to restrictions placed on use of NHS routinely collected data and current HRA & IRAS approval. Any application to use original anonymised dataset will require HRA Confidentiality Advisory Group approval. Data access point of contact.

## Acknowledgements

The authors thank The High End Computing facility at Lancaster University for providing the facilities required to conduct the inference.

