## supplementary material for "A Bayesian approach to identifying the role of hospital structure and staff interactions in nosocomial transmission of SARS-CoV-2"

### Hospital Network

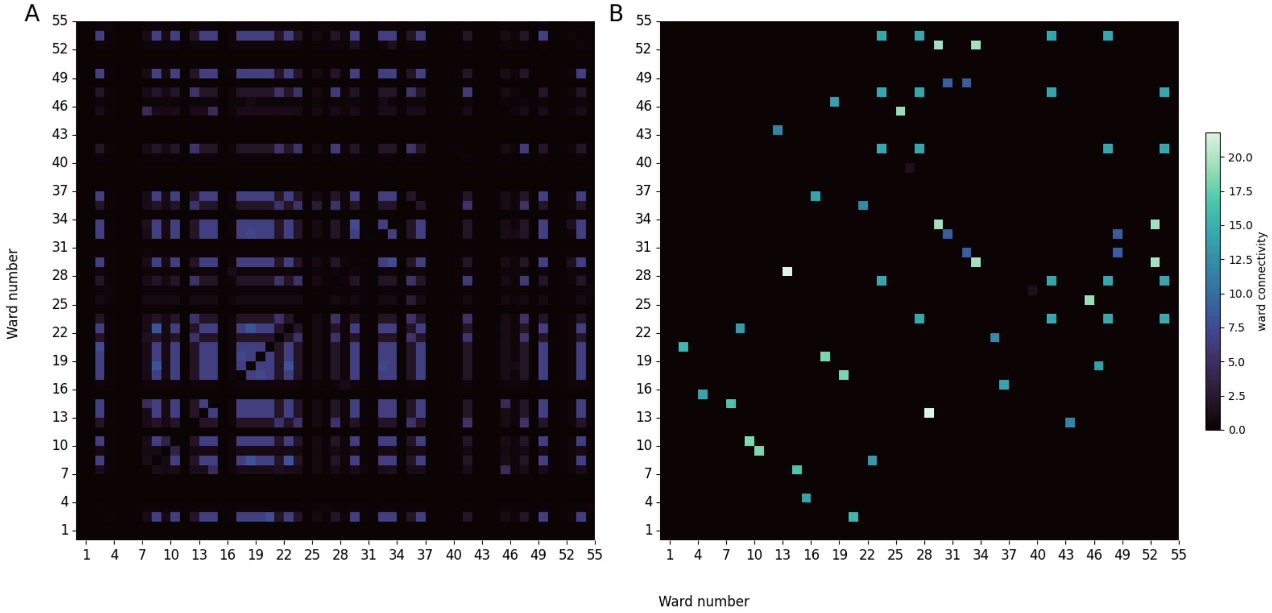


**Figure 1:** (A) Mean ward connectivity $C$ for the study period, weighted by staff assigned to multiple wards. (B) Mean ward proximity $W$, weighted by the number of staff assigned to each ward.

### MCMC algorithm

For each of the 11,000 iterations, we sample from the conditional posteriors for the transmission parameters, $\beta = (\beta_1, \beta_2, \beta_3, \beta_4, \beta_5)^\intercal$ in turn and approximately 10% of the unobserved state transition times, defined as the vector $U = (T_{SE}, T_{IR})$. We define the dimensions of $U$ as $dim(U) = [1, 320]$, and sample 33 $(320/10 + 1)$ event times per MCMC iteration.

**Algorithm 1:** Metropolis-within-Gibbs algorithm

**Input:** $\beta$, $U$ and tuning constants: $\sigma_\beta = (\sigma_{\beta_1}, \sigma_{\beta_2}, \sigma_{\beta_3}, \sigma_{\beta_4}, \sigma_{\beta_5})$, $\sigma_U$
**Output:** $\beta$, $U$
**Initialise:** $\beta^{(0)}$**,** $U^{(0)}$**,** $n = 0$**,** $x = 0$

**while** $n < 11,000$ **do**
  **for** $i$ *in 1 . . . 5* **do**
    $\beta^* = Normal(\beta_i, \sigma_{\beta_i})$
    **Compute**
      $\alpha(\beta_i, \beta^*) = 1 \ \wedge \frac{\pi(\beta^*|\beta_{-i},\, U)}{\pi(\beta_i|\beta_{-i},\, U)} \cdot \frac{\beta^*}{\beta_i}$
    **if** $Uniform(0, 1) < \alpha(\beta_i, \beta^*)$ **then**
      set $\beta_i = \beta^*$
    **end**
  **end**
  **while** $x < 33$ **do**
    Choose an event time to move, $U_j$, where $j = \text{randomInt}\big[0,\ dim(U)\big)$
    $U^* = Normal(U_j, \sigma_U)$
    **Compute**
      $\alpha(U_j, U^*) = 1 \ \wedge \frac{\pi(U^*|U_{-j},\, \beta)}{\pi(U_j|U_{-j},\, \beta)}$
    **if** $Uniform(0, 1) < \alpha(U_j, U^*)$ **then**
      set $U_j = U^*$
    **end**
    $x = x + 1$
  **end**
  $n = n + 1$
**end**
sssss

### Model Fit

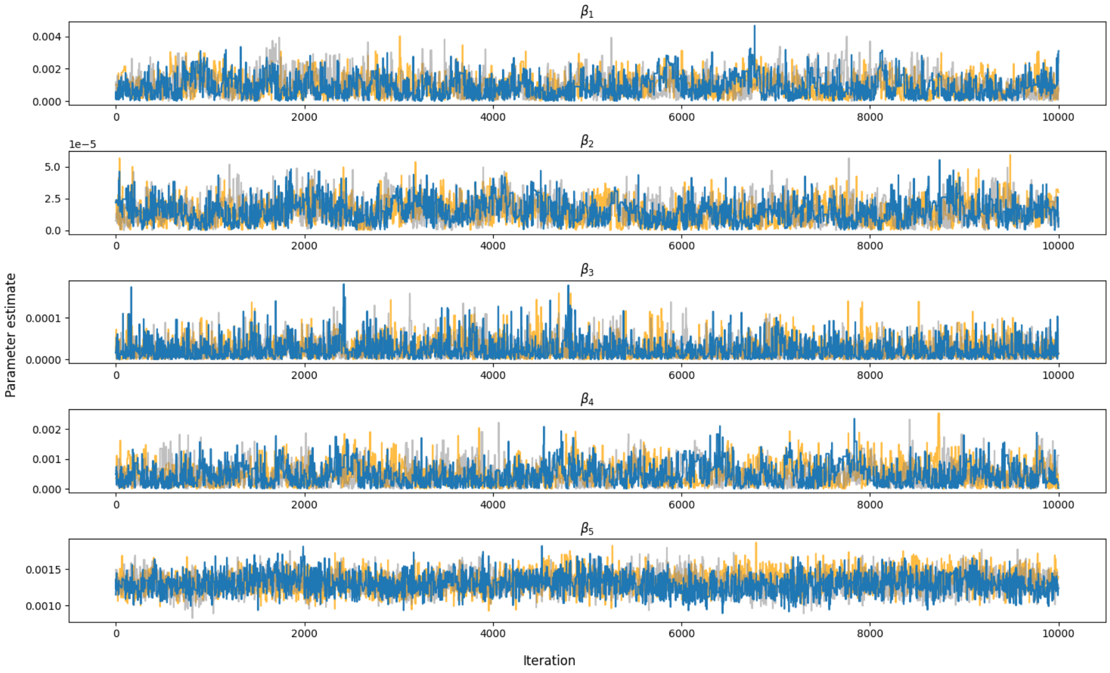


**Figure 2:** MCMC trace plots of three independent chains.

**Table 1:** Potential Scale Reduction Statistic.

| Parameter | Potential Scale Reduction Statistic |
|---|---:|
| $\beta_1$ | 1.02 |
| $\beta_2$ | 1.01 |
| $\beta_3$ | 1.00 |
| $\beta_4$ | 1.00 |
| $\beta_5$ | 1.04 |
